# Socio-virtual Broadcasting Platforms Contributing to Health Risks in sub-Saharan Africa: A Systematic Review Protocol

**DOI:** 10.1101/2025.09.29.25336884

**Authors:** Senzelokuhle. M. Nkabini, Lara. F. Croft, Lindiwe Nkabini-Anderson, Fezile Zuma

**Affiliations:** Faculty of Humanities, University of Pretoria. Hatfield Campus: Hatfield, Lynnwood Rd & Roper St, Pretoria, 0002. Gauteng province, South Africa; Faculty of Human Sciences, University of Campinas. Cidade Universitaria Zerefino Vaz, Barao Geraldo, Campinas-SP 13083-970, Brazil. (Visiting Academic); Clean Up Project,728 P Imfume Main Road, Amanzimtoti,4125 KwaZulu-Natal: South Africa

**Keywords:** Socio-virtual Broadcasting Platforms, Health Risks, sub-Saharan Africa, SSA

## Abstract

**Background:** The development and continuous advancement of the internet, has contributed to the significant progression of globalisation and international relations. This improvement has been spearheaded by the numerous sub-sections of social media that exist on the internet, the introduction of ‘influencer culture’, and the gradual evolution of continental, and regional economies. The socio-virtual interaction of the global community has ensured that both eccentric and conventional citizens, from either industrialised, economically progressing, or under-developed countries are able to coexist on interactive socialising platforms (social media). In 2021 4.87 billion people were using socio-virtual broadcasting platforms, and the number has increased to 5.56 billion in 2025. Within the sub-Saharan African (SSA) region, 45,4% (West Africa: 42.5%, Middle Africa: 33.6%, East Africa: 28.5%, & Southern Africa: 77,0%) of the 1.6 billion citizens also use these platforms. However, despite the shared commonalities amongst the viewers; the excessive financial benefits for the content creators & influencers; and the economic advancements for conglomerates, socio-virtual broadcasting platforms have deliberately and subconsciously contributed to the current existing global, continental and regional health risks. This is due to inadequate physical activity, unhealthy eating habits, the excessive consumption of alcohol & energy drinks, and disrupted sleeping patterns because of long hours spent on these platforms (*Virtually participating in Cookbangs* & *Mukbangs, Podcasting, Live-streaming* & *Gaming*). In SSA, the current existing NCD’s are cancer = 543,471 deaths, chronic respiratory disease (CRD)=178,908 deaths, cardiovascular disease (CVD)= 1 million deaths, diabetes=416,000 deaths, poor mental health issues (leading to suicide) =200,000 deaths, and chronic kidney disease (CKD)=108,185 deaths.

**Methods:** The primary research question that will guide this review is: How have socio-virtual broadcasting platforms contributed to the current existing health risks in SSA? The secondary research question is: What evidence exists regarding socio-virtual broadcasting platforms contributing to health risks in SSA? These research questions will assist in systematically scoping, mapping, and synthesising evidence of socio-virtual broadcasting platforms contributing to health risks in SSA. The following databases will be utilized to search for studies: PubMed, MEDLINE, ERIC, and Cochrane Reviews. The Preferred Reporting Items for Systematic and Meta Analyses (PRISMA) ScR flow chart/diagram presented in figure 1 will be utilized to summarize the study selection process. A data charting table will be used in order to document the extracted data. A narrative synthesis will be conducted, which will provide texts and tables in order to synthesize and discuss the data from the studies and the methods.

**Conclusion:** The commonness of content creators/influencers indulging in tobacco & synthetic nicotine products, alcoholic beverages, and psychoactive substances, whilst utilising socio-virtual broadcasting platforms (during *Cookbangs* & *Mukbangs, Vlogs, recording short-form content, Podcasting, Live-streaming* & *Gaming*) is alarming. Globally, substances such as alcohol (2,5 billion), tobacco (approximately 1,3 billion), and psychoactive drugs (approximately 500 million) are consumed at an astronomical rate. Hence, the continuous global increase in health risks and NCD’s. Regrettably, only 12% of countries throughout the globe have restricted the advertising of these products on the internet and socio-virtual broadcasting platforms. Moreover, the encouragement of casual sex/hook-up culture on numerous socio-virtual broadcasting platforms is problematic. The global escalation of sexually transmitted infections (STI’s) such as bacterial STI’s (chlamydia, gonorrhoea, syphilis), and viral STI’s (HIV, genital herpes, hepatitis, HPV) in October 2025 is currently 273 million (more than 1 million people are newly infected with STI’s each day). The proposed systematic review will generate findings pertaining to socio-virtual broadcasting platforms contributing to health risks in SSA. These findings will/can reveal the current existing literature gaps, and inform humanitarian organisations such as WHO, UNESCO, and UNICEF.

**Systematic reviews registration:** PROSPERO (CRD420251157784)

## Background

The development and continuous advancement of the internet, has contributed to the significant progression of globalisation and international relations. This improvement has been spearheaded by the numerous sub-sections of social media that exist on the internet, the introduction of ‘influencer culture’, and the gradual evolution of continental, and regional economies (1,2,3,4). The socio-virtual interaction of the global community has ensured that both eccentric and conventional citizens, from either industrialised, economically progressing, or under-developed countries are able to coexist on interactive socialising platforms (social media) (1,2,3,4). In 2021 4.87 billion people were using socio-virtual broadcasting platforms, and the number has increased to 5.56 billion in 2025 (8). Within the sub-Saharan African (SSA) region, 45,4% (West Africa: 42.5%, Middle Africa: 33.6%, East Africa: 28.5%, & Southern Africa: 77,0%) of the 1.6 billion citizens also use these platforms (8). The numerous subdivisions of social media have created individualistic, and preferential conclaves that are centred on the viewers or subscribers’ entertainment necessities (1,2,3,4). These conclaves range from *Cookbangs* & *Mukbangs* (videorecording the process of preparing, cooking and eating the food that has been cooked), *Vlogs* (videorecording one’s life on a daily or weekly basis), *Storytimes* (talking about an event or period), and various forms of short-form content (1,2,3,4). Moreover, *Podcasting*, *Live-streaming* & *Gaming* (communicating with viewers or fellow game players on a live session for an extended period) has become a new fad in the socio-virtual broadcasting community. However, despite the shared commonalities amongst the viewers; the excessive financial benefits for the content creators & influencers; and the economic advancements for conglomerates, socio-virtual broadcasting platforms have deliberately and subconsciously contributed to the current existing global, continental and regional health risks (1,2,3,4). In the SSA region, these platforms have aided in the upsurge of numerous health risks and other new diseases.

Health risks are lifestyle choices that lead to non-communicable diseases (NCD’s) or other negative health consequences that gradually affect humans (5,6,7). Other components such as age, and genetic predispositions can also be regarded as health risks (5,6,7). In SSA, the current existing NCD’s are cancer = 543,471 deaths, chronic respiratory disease (CRD)=178,908 deaths, cardiovascular disease (CVD)= 1 million deaths, diabetes=416,000 deaths, poor mental health issues (leading to suicide) =200,000 deaths, and chronic kidney disease (CKD)=108,185 deaths (5,6,7,14). The health risks that have contributed to these NCD’s range from a multitude of personal choices, individual substance induced traumatic experiences & societal customs (harmful alcohol consumption, tobacco use, vaping, unhealthy diets, & physical inactivity), coupled with environmental hazards (air pollution) (5,6,7). Moreover, numerous United Nations (UN) organisations such as, the World Health Organisation (WHO), the United Nations Inter-Agency Task Force on the Prevention and Control of NCD’s (UNIATF), United Nations Development Programme (UNDP), and United Nations Children’s Fund (UNICEF) are currently implementing strategies to decrease the upsurge of these NCD’s. However, due to the invention and continuous development of the internet, socio-virtual broadcasting platforms have fostered the continuous increase in the current existing NCD’s. This is due to inadequate physical activity, unhealthy eating habits, the excessive consumption of energy drinks, and disrupted sleeping patterns because of long hours spent on these platforms (*Virtually participating in Cookbangs* & *Mukbangs, Podcasting, Live-streaming* & *Gaming*) (12,13). In addition, cyberbullying on socio-virtual broadcasting platforms has led to health risks such as depression (resulting in suicide ideation), and addiction to attention has led to viewers partaking in physically risky behaviours because they are influenced by the insatiable need of obtaining ‘likes and views’ (9,10,11). Hence, the physical health, mental well-being and life expectancy of SSA citizens that utilise these platforms has started to decline.

Globally, socio-virtual broadcasting platforms have been used to promote unhealthy eating habits in low-income countries (LIC’s) (65%), and lower-middle-income countries (LMIC’s) (73%) (15). The numerous health violations and promotion of ‘deep-fried’ diets by the viewers and the content creators/influencers on these platforms has increased from 7% to 11% (17). In SSA, there are approximately 41 countries out of 49 that are defined as LIC’s and LMIC’s (16). Within these LIC’s and LMIC’s, 45% (29 million) of 13- to 19-year-old school children are currently overweight (15). In addition, the number of overweight children, adolescents, and adults in SSA LIC’s increased from 5 million to 19 million, and in LMIC’s it spiked from 23 million to 76 million (15). Food products such as soft drinks, sugary diets, and salty processed products in SSA are consumed at an excessively high rate (West & Central Africa =71%; East & Southern Africa = 98%) than other regions or continents (15). These unhealthy eating habits coupled with the lack of physical activity, have slowly created a region that is inhabited by overweight and obese citizens. This crisis might also be exacerbated by 85% of SSA countries lacking in fruits & vegetables, whilst 37% of influencers in SSA LIC’s and LMIC’s continue to steadily promote unhealthy eating habits, and a physically inactive lifestyle on socio-virtual broadcasting platforms (15). Thus, the need for global, continental, and regional programmes coupled with implementable strategies to counter this issue.

The *Teens, Screens and Mental Health* survey was initiated in 2021-2022 by the WHO Europe regional office. This survey focused on tracking, monitoring, and assessing self-reported risky behaviours; health outcomes regarding adolescent socio-virtual broadcasting platforms & gaming, and the gradual development of addiction to these platforms (17). The countries and regions that took part in this survey were from Europe, central Asia, and Canada. The results from the survey encouraged countries and regions to: (i) consider their modus operandi on regulations and access, to digital technologies, (ii) assess socio-virtual broadcasting platforms & gaming regulatory frameworks that have been drafted, approved, and set for implementation, in order to ensure that content which is posted and advertised on these platforms adheres to the digital technologies, parental and educational guidelines, (iii) ensure that socio-virtual broadcasting platforms & gaming sites are utilised in a healthy manner that encourages physical activity, individual self-mental health assessment, and healthy eating habits during the usage of these platforms (17). In spite of the investigation promoting robust frameworks that focus on a self-monitored healthy utilisation of socio-virtual broadcasting platforms, this survey was aligned with countries and regions that have advanced economies (high developed incomes with advanced innovations), and the necessary resources. Moreover, the LIC’s and LMIC’s in the SSA region are the most impacted by the promotion of unhealthy eating habits, the marketing of pseudoscience, and the publicising of untested and unapproved medical treatments/home-remedies on socio-virtual broadcasting platforms (15,18,19). The censorship of false medical treatments/home-remedies in both LIC’s and LMIC’s in SSA is currently at 0%, as indicated by a UNICEF study published in 2025 (15). Hence, the vital need for WHO surveys such as *Teens, Screens and Mental Health* to also be inclusive of countries that have developing (LIC’s), and emerging economies (LMIC’s) such as those in the SSA region.

SDG 3: *Good Health and Well-being* is part of the Sustainable Development Goals (SDG’s) that were formulated by the UN to address the societal, economic, health, and educational challenges that are a big impact globally, continentally, and regionally (26). The unhealthy lifestyle habits, pseudoscience, and unapproved medical treatments promoted on socio-virtual broadcasting platforms, form part of the challenges that should be addressed by SDG 3. A vast majority of content creators and influencers on these platforms promote unhealthy: food choices, excessive alcohol consumption, and smoking habits (20,22,23). These habits contribute to the prominent leading causes of death (cancer, CRD, CVD, diabetes, poor mental health issues & CKD) in SSA LIC’s and LMIC’s, coupled with vision & hearing loss, weaker bones, digestive issues, erectile dysfunction, and ectopic pregnancies (21,24,25). Socio-virtual broadcasting platforms are vital to ensuring the reduction, prevention and treatment of NCD’s, health risks, and other neglected diseases. Hence, it is crucial to ensure that these extremely addictive, and influential platforms are utilised by adolescents and adults in a trustworthy, informative, and beneficial manner.

The primary research question that will guide this review is: How have socio-virtual broadcasting platforms contributed to the current existing health risks in SSA? The secondary research question is: What evidence exists regarding socio-virtual broadcasting platforms contributing to health risks in SSA? These research questions will assist in systematically scoping, mapping, and synthesising evidence of socio-virtual broadcasting platforms contributing to health risks in SSA. A preliminary database search has been conducted in order to prevent the duplication of previous reviews. Databases such as PubMed, MEDLINE, ERIC (Education Resources Information Center), and Cochrane Reviews were utilised to conduct this preliminary search. The research team (SMN, LC, LN-A & FZ) has no knowledge of previous systematic reviews being conducted on socio-virtual broadcasting platforms contributing to health risks in SSA. Thus, further research is required on this topic and the results from this systematic review will assist organisations such as WHO, United Nations Educational, Scientific and Cultural Organization (UNESCO), and UNICEF.

## Methods

The primary aim of this systematic review is to scope, map out, and synthesise evidence of socio-virtual broadcasting platforms contributing to health risks in SSA from existing literature. This systematic review protocol is registered on the International Prospective Register for systematic reviews. All forms of studies, grey literature and peer-reviewed journal articles focusing on socio-virtual broadcasting platforms, and health risks in SSA will be sourced. The primary research question that will guide this review is: How have socio-virtual broadcasting platforms contributed to the current existing health risks in SSA? The secondary research questions are: What evidence exists regarding socio-virtual broadcasting platforms contributing to health risks in SSA? The P.I.Co (population, interest, context) framework was utilized to address the eligibility, and adequateness of the primary and secondary research questions. This framework (P.I.Co) has been adapted from the P.I.C.O framework, which is used for quantitative systematic reviews (27). However, due to this systematic review focusing on socio-virtual broadcasting platforms contributing to health risks in SSA, which are qualitative components, the P.I.Co framework will be utilised (27).

### P.I.Co (Population, Interest, Context) framework

**Population:** Socio-virtual broadcasting platforms are *Twitch*, *Blue Sky*, *YouTube*, *Facebook Live*, *TikTok*, *X* (formerly known as *Twitter*), *Snapchat*, *Threads*, *WhatsApp*, *Roblox* and *Instagram*. These platforms allow for real-time streaming and engagement with large audiences for various purposes, such as gaming or commerce. Moreover, the users of these platforms in SSA, range from adolescents, youths, adults, and older members of society. The populations basic, moderate, and elevated knowledge of socio-virtual broadcasting platforms is dependent on the SSA country’s economic status/ranking (28,29).

Age: 13+ years old is the average age that is allowed to use socio-virtual broadcasting platforms (15).

**Interest:** Health risks in SSA, are exacerbated by the deception, propaganda, and the uncouthness exemplified by content creators or influencers on socio-virtual broadcasting platforms (30).

**Context:** SSA is a region within the African continent that has 49 countries (31). **Sources of evidence:** All studies, grey and empirical literature containing evidence on socio-virtual broadcasting platforms contributing to health risks in SSA.

**Publication Year Range:** 01/01/2021-31/10/2025 **Language:** No language restriction

### Information sources and search strategy

All searches will be conducted by SMN, LC, LN-A and FZ for studies and study designs published in peer-reviewed journals, grey literature, published and unpublished dissertations, case studies, reviews, essays, theses and symposium abstracts. The following databases will be utilized to search for studies: PubMed, MEDLINE, ERIC, and Cochrane Reviews. The search strategy will use the Boolean term ‘OR’ to separate words or search terms. The following search terms will be utilised: socio-virtual OR broadcasting OR platforms OR contributing OR health risks OR sub-Saharan Africa OR SSA. A preliminary data base search was conducted by the research team using the search terms, and the results are presented in Table 1. The preliminary search produced all studies, qualitative and quantitative full-text grey literature and peer-reviewed literature with no language restrictions, and published within the search time-line from 01January 2021 till 31 October 2025.

**Table 1:**
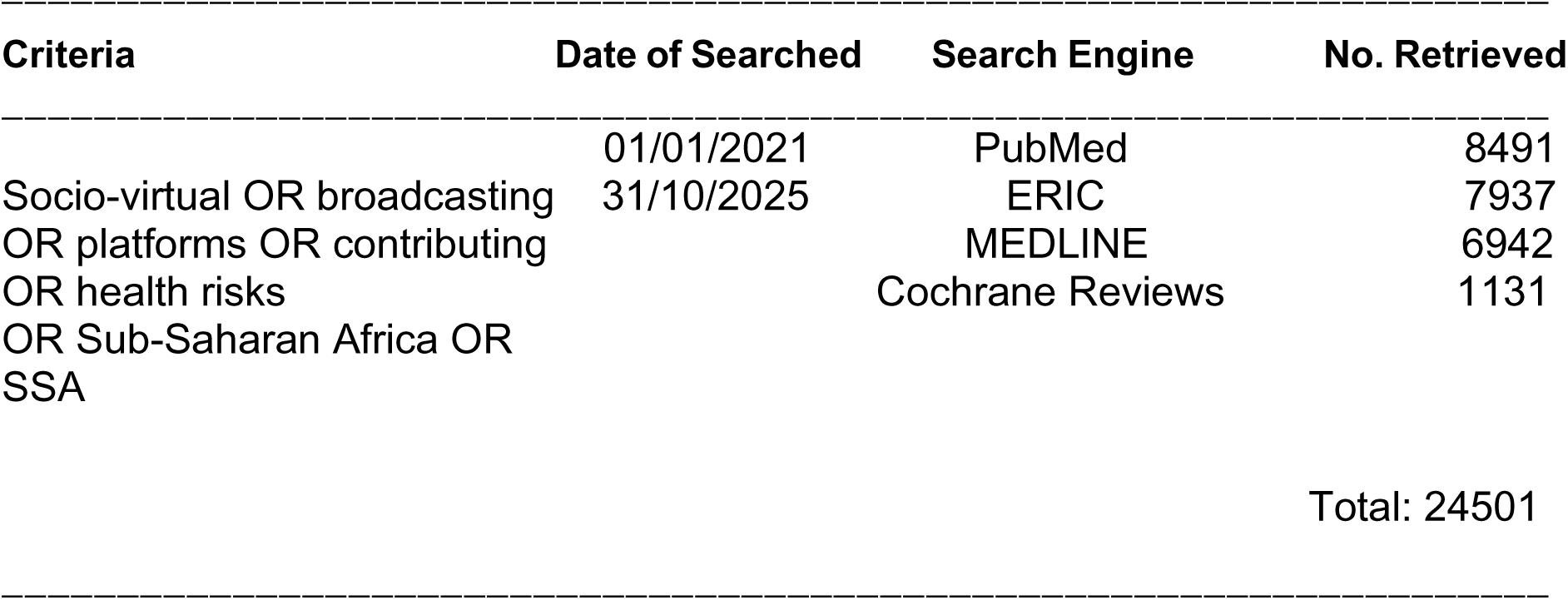
Database search.

**Table 2:** Data charting table.

|  |
| --- |
| Author and year of publication |
| Article title |
| Study aim |
| Country |
| Study design/methodology |
| Study setting |
| Study population |
| Duration of the study |
| Socio-virtual broadcasting platforms contributing to health risks in SSA |
| Population age |
| Relevant/significant findings |
| Key conclusions of article |
| Notes |

### Inclusion and exclusion criteria

The selection of eligible studies will be based on the title (socio-virtual broadcasting platforms contributing to health risks in SSA), abstract, setting (SSA), study population (socio-virtual broadcasting platform users) and the findings in full-text studies, books, grey literature and journal articles relating to socio-virtual broadcasting platforms, and health risks in SSA. All database searches will be conducted on a weekly basis by SMN, LC, LN-A and FZ to ensure new literature is included. Only studies that adhere to the following criteria will be included: (i) socio-virtual broadcasting platforms contributing to health risks in SSA, (ii) qualitative and quantitative study designs iii) studies and articles published from 2021 to 2025 and, (iii) only studies, books, grey literature or journal articles that are restricted to human ages 13+ years old, full-text, have references/citations and have been peer-reviewed. Studies that are: (i) published before 2021, will be excluded.

### Data management and study selection

The framework, enhancements to the framework and guidelines provided by Arksey and O’Malley’s (32), Levac *et al* (33), Daudt *et al* (34), and Johanna Briggs (35) will guide this review. The Preferred Reporting Items for Systematic and Meta Analyses (PRISMA) ScR flow chart/diagram presented in figure 1 will be utilized to summarize the study selection process (36,37). The authors and the two research assistants will ensure that all retrieved literature will be exported and saved to an Endnote 21.3 library folder, in order to enable the study to be reproduced for a second time at a later stage. Moreover, this process will allow SMN, LC, LN-A and FZ to create separate libraries for each database, in order to import references, remove duplicates, and organize the findings.

**Figure 1:**
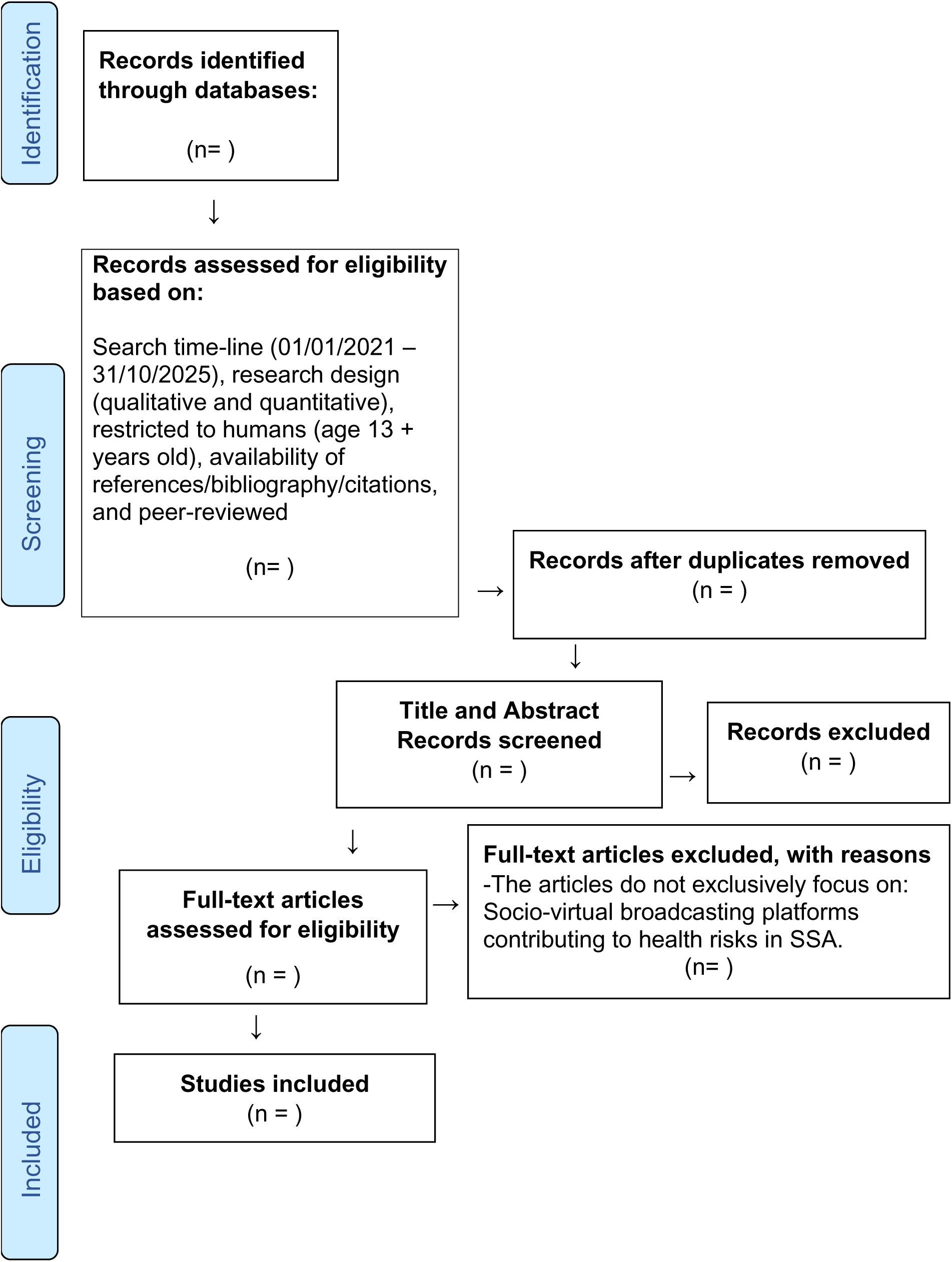
The Preferred Reporting Items for Systematic and Meta Analyses (PRISMA) ScR flow chart/diagram

### Data extraction

A data charting table will be used in order to document the extracted data. Studies that meet the inclusion criteria will be included on the charting form. The use of the NVivo data analysis & narrative summarizing software, and Braun & Clarke’s (38) thematic framework will guide the qualitative analysis. The data extraction form will be updated by SMN, LC, LN-A and FZ on a weekly basis to guarantee accurateness.

### Risk of bias assessment

Studies or articles that only feature few countries that are located in the SSA region will not be included, because the research results from these studies do not represent SSA as a whole. In order to avoid biasness the review will utilise the mixed-method appraisal tool (MMAT) version 2018 to appraise the quality of all included evidence (39). SMN, LC, LN-A & FZ will be responsible for assigning ratings of 100% for high average sources, 50% average and 25% for low-quality articles.

### Discrepancies between the protocol and the systematic review

Discrepancies between the protocol, the actual review and the reasons and consequences thereof will be reported in the final report.

### Data synthesis

A narrative synthesis will be conducted, which will provide texts and tables in order to synthesize and discuss the data from the studies and the methods, as previously described in the data extraction section.

## Results

The results of this systematic review will be disseminated through publication in peer-reviewed scientific journals. The data will also be made available to humanitarian organisations such as WHO, UNESCO, and UNICEF. The findings from this study will inform the future research projects that these organisations embark on.

### Ethical approval

The systematic review will not involve human or animal subjects. Thus, ethical approval is not necessary for the systematic review.

## Discussion

The commonness of content creators/influencers indulging in tobacco & synthetic nicotine products, alcoholic beverages, and psychoactive substances, whilst utilising socio-virtual broadcasting platforms (during *Cookbangs* & *Mukbangs, Vlogs, recording short-form content, Podcasting, Live-streaming* & *Gaming*) is alarming. Globally, substances such as alcohol (2,5 billion), tobacco (approximately 1,3 billion), and psychoactive drugs (approximately 500 million) are consumed at an astronomical rate (40). Hence, the continuous global increase in health risks and NCD’s. Regrettably, only 12% of countries throughout the globe have restricted the advertising of these products on the internet and socio-virtual broadcasting platforms (40). Moreover, the encouragement of casual sex/hook-up culture on numerous socio-virtual broadcasting platforms is also problematic. The global escalation of sexually transmitted infections (STI’s) such as bacterial STI’s (chlamydia, gonorrhoea, syphilis), and viral STI’s (HIV, genital herpes, hepatitis, HPV) in October 2025 is currently 273 million (more than 1 million people are newly infected with STI’s each day) (41). Additionally, within the global context, socio-virtual broadcasting platforms have contributed to the high levels of cyberbullying, which is also enacted through the sharing of sexually explicit A.I. generated images and videos. 16% of adolescents in Europe, central Asia and Canada reported that they had been cyberbullied at least once or twice on a socio-virtual broadcasting platform (42). WHO studies and surveys such as *Teens, Screens and Mental Health*; the *Global status report on alcohol and health and treatment of substance use disorders*; the *Global health sector strategies on, respectively, HIV, viral hepatitis and sexually transmitted infections for the period 2022-2030*; and *A focus on adolescent peer violence and bullying in Europe, central Asia and Canada* do not include SSA regions in their studies/surveys and some do not even consider the African continent as a research sample. Thus, the proposed systematic review will generate findings pertaining to socio-virtual broadcasting platforms contributing to health risks in SSA. These findings will/can reveal the current existing literature gaps, and inform humanitarian organisations such as WHO, UNESCO, and UNICEF on this critical issue.

## Data Availability

All data produced in the present work are contained in the manuscript

## Acknowledgments

The authors express their gratitude to: the Faculty of Humanities at the University of Pretoria, and the Faculty of Human Sciences at the University of Campinas.

## Authors contributions

SMN, LC, LN-A and FZ conducted the preliminary database search for the protocol, conceptualised, drafted, edited, reviewed, sent out the draft protocol manuscript for constructive feedback, and approved the final manuscript.

## Ethics approval and consent to participate

Not applicable.

## Consent for publication

Not applicable.

## Availability of data material

The required data is included in the manuscript.

## Abbreviations

A.I.: Artificial Intelligence
CRD: Chronic Respiratory Disease
CVD: Cardiovascular Disease
CKD: Chronic Kidney Disease
ERIC: Education Resources Information Center
HIV: Human Immunodeficiency Virus
HPV: Human Papillomavirus
LIC: Low-Income-Country
LMIC: Lower-Middle-Income-Country
MMAT: Mixed-Method Appraisal Tool
NCD: Non-Communicable Disease
P.I.Co: Population. Interest. Context
PRISMA: Preferred Reporting Items for Systematic and Meta Analyses
SSA: sub-Saharan Africa
SDG: Sustainable Development Goals
STI: Sexually Transmitted Infections
UN: United Nations
UNIATF: United Nations Inter-Agency Task Force on the Prevention and Control of NCD’s
UNDP: United Nations Development Programme
UNICEF: United Nations Children’s Fund
UNESCO: United Nations Educational, Scientific and Cultural Organization
WHO: World Health Organisation

---

|  |  |  | Page |
| --- | --- | --- | --- |
| Section and | Item No | Checklist item topic |  |
| **ADMINISTRATIVE INFORMATION** |  |  |  |
| Title: |  |  |  |
| Identification | 1a | Identify the report as a protocol of a systematic review | Page 1 |
| Update | 1b | If the protocol is for an update of a previous systematic review, identify as such | N/A |
| Registration | 2 | If registered, provide the name of the registry (such as PROSPERO) and registration number | Page 3 |
| Authors: Contact |  |  |  |
|  | 3a | Provide name, institutional affiliation, e-mail address of all protocol authors; provide physical mailing address of corresponding author | Title Page<br>Page 1 |
| Contributions | 3b | Describe contributions of protocol authors and identify the guarantor of the review | Page 14 |
| Amendments | 4 | If the protocol represents an amendment of a previously completed or published protocol, identify as such and list changes; otherwise, state plan for documenting important protocol amendments | N/A |
| Support: Sources |  |  |  |
|  | 5a | Indicate sources of financial or other support for the review | Pages 15 |
| Sponsor | 5b | Provide name for the review funder and/or sponsor | N/A |
| Role of sponsor or funder | 5c | Describe roles of funder(s), sponsor(s), and/or institution(s), if any, in developing the protocol | N/A |
| **INTRODUCTION** |  |  |  |
| Rationale | 6 | Describe the rationale for the review in the context of what is already known | Pages 4, 5, 6 & 7 |
| Objectives | 7 | Provide an explicit statement of the question(s) the review will address with reference to participants, interventions, comparators, and outcomes (PICO)- P.I.Co. Systematic review | Page 8 |
| **METHODS** |  |  |  |
| Eligibility criteria | 8 | Specify the study characteristics (such as PICO, study design, setting, time frame) and report characteristics (such as years considered, language, publication status) to be used as criteria for eligibility for the review- P.I.Co. Systematic review | Pages 8 & 9 |
| Information sources | 9 | Describe all intended information sources (such as electronic databases, contact with study authors, trial registers or other grey literature sources) with planned dates of coverage | Page 9 |
| Search strategy | 10 | Present draft of search strategy to be used for at least one electronic database, including planned limits, such that it could be repeated | Page 9 |
| Study records: |  |  |  |
| Data management | 11a | Describe the mechanism(s) that will be used to manage records and data throughout the review | Page 10 |
| Selection process | 11b | State the process that will be used for selecting studies (such as two independent reviewers) through each phase of the review (that is, screening, eligibility and inclusion in meta-analysis) | Pages 10, 11 & 12 |
| Data collection process | 11c | Describe planned method of extracting data from reports (such as piloting forms, done independently, in duplicate), any processes for obtaining and confirming data from investigators | Page 12 |
| Data items | 12 | List and define all variables for which data will be sought (such as PICO items, funding sources), any pre-planned data assumptions and simplifications | Pages 14 & 15 |
| Outcomes and prioritization | 13 | List and define all outcomes for which data will be sought, including prioritization of main and additional outcomes, with rationale | Pages 13 & 14 |
| Risk of bias in individual studies | 14 | Describe anticipated methods for assessing risk of bias of individual studies, including whether this will be done at the outcome or study level, or both; state how this information will be used in data synthesis | Pages 12 & 13 |
| Data synthesis | 15a | Describe criteria under which study data will be qualitatively synthesised | Page 13 |
| | 15b | If data are appropriate for quantitative synthesis, describe planned summary measures, methods of handling data and methods of combining data from studies, including any planned exploration of consistency (such as $I^2$, Kendall's $\tau$) | N/A |
|  | 15c | Describe any proposed additional analyses (such as sensitivity or subgroup analyses, meta-regression) | N/A |
|  | 15d | If quantitative synthesis is not appropriate, describe the type of summary planned | N/A |
| Meta-bias(es) | 16 | Specify any planned assessment of meta-bias(es) (such as publication bias across studies, selective reporting within studies) | Pages 12 |
| Confidence in cumulative evidence | 17 | Describe how the strength of the body of evidence will be assessed (such as GRADE) | Page 12 & 13 |

**\* It is strongly recommended that this checklist be read in conjunction with the PRISMA-P Explanation and Elaboration (cite when available) for important clarification on the items. Amendments to a review protocol should be tracked and dated. The copyright for PRISMA-P (including checklist) is held by the PRISMA-P Group and is distributed under a Creative Commons Attribution Licence 4.0. From: Shamseer L, Moher D, Clarke M, Ghera D, Liberati A, Petticrew M, Shekelle P, Stewart L, PRISMA-P Group. Preferred reporting items for systematic review and meta-analysis protocols (PRISMA-P) 2015: elaboration and explanation. *BMJ*. 2015 Jan 2;349(jan02 1):g7647.**

